# Underlying Pressures that Black Mothers and Their Children Face: A Qualitative Assessment on the Effects of Racism/Discrimination and the COVID 19 Pandemic

**DOI:** 10.1101/2023.02.15.23286011

**Authors:** Denisia N. Thomas, Ashley Rizzieri, Mayra Rodriguez

## Abstract

**Background:** Poverty, racism, discrimination, inadequate access to healthcare, and personal, everyday stressors lead to poor health outcomes, especially in African American families in the south. There is limited data on how these stressors are absorbed between the mother and child dyad.

**Objective:** To assess the effects of racism, everyday stressors (i.e. motherhood), and the COVID pandemic on African American/Black mothers and their children.

**Methods:** Utilizing the Health Belief Model, a survey was developed to assess mother-child stressors relating to three different constructs: racism/discrimination, pandemic/covid-19, and parenting. We interviewed seven black mothers and their children (aged 4-10yo). The families were recruited from a pediatric office in the rural city of Alexander City, Alabama. Interviews took place in an intimate setting and lasted for 1.5-2 hours. Medical students conducted, recorded, and transcribed each interview. The interview assessed the association between the COVID-19 pandemic, personal traumatic events, and racism and discrimination in their everyday lives.

**Results:** Through qualitative analysis; racism, daily activities, and the COVID-19 pandemic were demonstrated to be significant stressors for the mothers. Knowledge, school/work, actions, emotions, and seriousness/susceptibility displayed stressors not only in the mom as one would expect, but in the children as well. Using the resilience model, we assessed adversity, coping strategies, and self-efficacy. As one might expect, each situation caused a different level of anxiety; however, the coping strategies varied. Some moms took to smoking to cope with it while others chose suppression. The children’s coping ranged from inconsolable crying and using outlets such as phones to cope.

**Conclusion:** Ultimately, our qualitative approach saw an association between the pandemic and discrimination. Mothers often felt the need to shield children from the emotions attached to discrimination, and ultimately were unable to. There is a need to explore resilience and assess these stressors and changes in perception over time.

## INTRODUCTION

Witnessing the Black Lives Matter protests and the unproportionable deaths of African Americans due to the COVID-19 pandemic drove the inspiration for this study. Poverty, racism, discrimination, inadequate access to healthcare, and personal, everyday stressors such as parenting lead to poor health outcomes, especially in African American families in the south. In the southern United States, influences of environmental stressors, such as poverty, racism, lack of medical care access, and personal stressors such as parenting or mental health, decrease the health outcomes in families [1-5]. In 2020, the COVID pandemic hit and disproportionately affected black families, killing more black Americans than their respective population proportional [9]. Along with the COVID pandemic, racism, and discrimination took center stage leaving Black/African Americans in the center of the storm. Black families all over the country expressed their anger, fear, and sadness with the current state of racism in America. The pandemic and current racism in America place a huge burden of stress on black families and their loved ones, adding to the already heavy load of stressors families face in underserved Alabama. Currently, there is limited data on how these accumulated stressors affect the Black mother and child relationship. This study aims to investigate the effects of the COVID-19 pandemic and racism on black mothers and their young children using in-depth interviews.

## METHODS

We used a semi-structured in-depth interview approach to gather data from seven Black/African American families from pediatric clinics in Alexander City.

One-on-one interviews were conducted in an intimate setting with the mothers and children, separately. The in-depth interviews addressed three different stressors, racism/discrimination, covid-19 pandemic, and parenting. Each of these stressors was assessed using the Health Belief Model as a framework with the following constructs: knowledge, school/work, action, emotions, and seriousness/susceptibility. Each of the questions were open-ended and trained staff were asked to allow time for a response. Interviewers were encouraged to ask, “tell me more” when there were opportunities to do so. Although we asked questions about covid-19 and the vaccine, interviewers avoided using jargon language. Interviewers were trained to be active listeners and treat their participants with respect. In the event a participant became upset, the interviewers were trained to allow time for the participant to take a moment to collect themselves or discontinue the interview. While the mothers were being interviewed, the children were in the room either interviewing with another staff member or playing/coloring waiting their turn to be interviewed.

Second-year nursing and medical students were trained in delivering the semi-structured in-depth interviews for mom and child. The semi-structured approach was beneficial when working with small children as it was difficult to maintain their attention during the interview. Interviews lasted approximately 1.5 hours per family. Data was recorded and transcribed by trained medical students. After transcription, the team met to discuss each of the transcript files where inaudible data was discussed to determine the accuracy of the file.

## RESULTS

### Pandemic

#### Knowledge

Almost all mothers know that Covid is a virus and could be prevented from standing six feet apart and wearing a mask as per CDC guidelines. Some mothers were able to expand on this stating that symptoms of the virus were “comparable to the common cold” and “extreme cases could lead to pneumonia”. All children knew about Covid-19 from either their parents or through television, but only some were able to explain how it was spread via “not wearing a mask” or “not standing six feet apart”. All children could explain that germs in general were shared with a lack of hand washing and distancing. Knowledge of the vaccine varied amongst mothers and children alike. Some mothers admitted to not knowing much about the vaccine and not being vaccinated themselves. Some admitted to needing to do more research on the vaccine because their knowledge was limited to personal opinions online. One mother expressed her initial hesitations about the vaccine and how she thought “it was gonna kill us”. Most children’s knowledge of the vaccine was limited. They heard of a vaccine either through school or alternative media outlets. Some children heard about the vaccine “hurting”.

#### Susceptibility/Seriousness

Moms who didn’t know anyone affected by the virus were still fearful of the hospitalizations and had anxiety discussing the virus. One mom wasn’t afraid of contracting the virus until she endured it herself. Another mom said she thought people pick and choose when to be serious about covid”, explaining that although she knew everyone was vulnerable, she would be more worried if someone in her family contracted the virus. Many moms believed that their children didn’t fully grasp the severity of Covid. Many of the children didn’t know anyone personally with the virus, didn’t believe they were susceptible to getting the virus, but did agree that it would “ be bad to be sick” and described it as a “10/10 of severity” if they were to get it. Many believed that other students and teachers at their school didn’t take the mask mandate or social distancing seriously.

#### School Work

All families, except for one who attended online, participated in school in person. Although the classrooms varied among the children, completing schoolwork was not a problem. The children attending school online did admit to “things changing. Hating school because it was too much work through virtual”. All children explained how they preferred to be in person during school. Those who were online missed their friends, the gym, and recess. Those who attended in person said things like “I like seeing my friends” and “I like when we get to go on the playground”.

#### Emotions

The gravity of the pandemic varied amongst the mothers, but all could admit to a degree of anxiety surrounding the virus. Mothers admit to facing adversity, “it wiped out all my residents and my sister passed away”. They express fear of what would happen to them if they were to get Covid themselves, “I’m always scared and don’t know what would happen if we were in that predicament”. Moms are not only stressed by getting the disease but also what it means to be absent from their jobs, “I’m working part-time as it is, and I can’t afford to not be at work”. Mothers don’t think the pandemic is something their children right now have to worry about, “I am happy he doesn’t worry and he doesn’t have time to worry”.

Children’s perceptions of the pandemic are that they are scared. Some expressed “I don’t want to get it, I just wanna stay away from people who have it. It feels nasty and yucky”. Some children are more worried about how it limits them in school and in friendships. One child admitted to feeling sad at times when she “wanted a hug from a friend but didn’t know if they had the virus”. Other children were annoyed when they had to wear their masks when it was hot outside or uncomfortable.

#### Action

Every mother and child admitted to wearing a mask, frequent hand washing, and social distancing. If they knew someone was in contact with the virus, then they would quarantine as per CDC guidelines.

### Racism

#### Knowledge

When asked what racism meant to mothers we received responses such as “when someone points out imperfections and singles them out for the way they are”, “hating another race of people”, and “people not wanting others to have the same status”. Many mothers have met and currently know racist people. When children were asked what they knew about racism, the responses were “when someone sees white vs. black”, “someone judges you for the way you look”, and some children didn’t know how to describe what racism was but heard about it through parents, school, or local news stations. Some children were aware of the recent protests going on in our nation surrounding racism.

#### Susceptibility/Seriousness

All mothers had experienced racism to some degree during their lives and believed they were generally highly susceptible to that form of discrimination. Many children deny experiencing racism within their schools, friendships, and community.

#### Emotion

Mothers and children both explained that discussing racism with friends, family, and in school made them uneasy, uncomfortable, sad, or mad. One mom explained how she faced an incidence of racism within the military and the conflict ran through her mind over again until the issue was resolved. Mothers who have experienced outward signs of racism in the past were worried that they will experience it again within their community and wanted to shelter their children from being victimized. One mom stated, “I want my child to be a child and don’t want him to worry about that”, explaining that she wants him to enjoy being a kid. Moms feel that they will have to raise their children in a particular way in the future. One mom describes how she feels raising her sons, “I have to raise them as black men, I have to teach them how to be strong and that’s hard”. She further elaborated on how she will have to instill resilience in her sons in light of current events surrounding police brutality.

Children admit to seeing protests on TV and seeing their parent’s reactions. One child compared the protests to a scary movie. Many children had not experienced racism within their school or friend groups.

#### Action

Some mothers have not outwardly had a discussion with their children about racism yet, but many have had discussions on responding to a bully. Advice given included, “walking away”, “not getting involved”, and “don’t retaliate”. Some moms educated their children through the news on TV and used it as a platform to explain how some members of society discriminate based on skin color and race. When children were asked what they would do if they experienced another being racist towards them, they all resulted in telling someone, most likely a principal of the school.

## DISCUSSION

Historically, American systems have excluded Black women from support and resources conferred on white women, an inequity exemplified by the welfare system (10). The participants agreed they were susceptible to covid, but the perceived severity of the virus varied based on individual experiences of knowing someone personally who was sick. Regardless, all participants were adherent to the mask mandate but wary of the vaccination. This can be attributed to a possible lack of knowledge of how the vaccine was made, the timing of the vaccine, and exposure to public opinions of fear and distrust in the vaccination in general. One recent study found that out of 963 Alabamians, 44.5% were unlikely or highly unlikely to get vaccinated. Furthermore, healthcare providers were the most trusted regardless of vaccination status, however those who were unlikely to be vaccinated trusted social media and their local news more (11). Covid-19 has placed undue stress on families to adjust their daily routines. Some children were forced to transition to a virtual classroom that allowed them to continue their studies but isolated them from their friends and denied them the opportunity to enjoy their favorite activities at school like recess. Some mothers were worried about how they would financially support their families due to layoffs from the pandemic. According to a recent executive labor summary in Alabama, the pandemic disproportionately affected women and persons of color more (12). Through the seven families interviewed we are already seeing a clear effect that Covid has on financial and emotional stability.

Mothers were motivated to protect their children from racism and social concerns. Many wanted them to enjoy being children and not have to worry about some of the harsh realities. It was, however, clear, that many would eventually have the discussion in the future. Even if not prompted directly by the parents, many were not hesitant to introduce concepts of racial injustice to their children through the exposure of recent events, including national protests against police brutality. Fortunately, many children interviewed denied experiencing outward signs of racism at school but had a plan for if they did, which usually involved telling an adult.

Apart from data pending, there are a few limitations to the results that possibly inhibited us from grasping a full picture of how stress from covid/racism affects coping behavior. The small sample size of seven moms and their children limits the variety of life events experienced. Some children were subjected to virtual classrooms while others still maintained in-person lessons at school. Within the responses from participants, we noticed a difference in the amount of schoolwork, exposure to play, and exposure to friendships based on the type of learning environment children were subjected to. One could imagine that the children who were exposed to a virtual classroom didn’t have the same coping mechanisms available as those attending in person.

In addition, bias could have been introduced within data collection due to different interviewers asking questions. Not all questions could have been asked the same way for each family. Not all families could have felt open to discussing these sensitive topics with each interviewer based on comfortability. These factors would most certainly affect the quality of answers given at the time.

There is a possibility that leading bias was introduced into the study as well due to redirecting children in each interview. The time required to complete each survey superseded the attention span of some participants. It led some interviewers to rephrase open-ended questions like, “how is your school work” to closed-ended questions like “you do well in school, right”?

The interplay between the pandemic and racism experienced by the families will have to be further analyzed upon completion of the quantitative arm of this study to see how it affects coping behaviors. Some moms admitted to crying, their children lashing out to get what they want, and moms picking up habits like smoking when faced with chronic stress in general. The qualitative arm examined in this study allows us to peek into some of the perceptions families hold but doesn’t give us a complete picture.

## CONCLUSIONS

As hypothesized, mothers followed CDC precautions and taught their children to prevent contracting Covid. In addition to mothers teaching their children how to be safe, school, and social media played a role in influencing mask behavior and social distancing as well. Many children were not convinced they were susceptible to covid but agreed it would be bad if they were sick with this virus. In addition, children were bothered by the limited playtime with friends at school and sometimes inconvenienced by the mask mandate when it was hot outside.

Many of the moms had been affected by racism in their lives but had not had a discussion with their children on how it presents and what to do if they themselves experienced it. Some children had an idea of what racism was and how they would respond to someone bullying them but had not admitted to experiencing it in their daily lives at school. To truly complete the picture of how covid and racism affect both mother and child, the stress they endure, and subsequent coping mechanisms we will need to finish the quantitative arm of this study.

## Data Availability

All relevant data are within the manuscript and its Supporting Information files.

## ACKNOWLEDGEMENTS

We would like to extend a special thank you to the following people that helped who participated in the development and execution of this project.

1. David Stephen, DO: Assistance with the quantitative arm of this study.
2. Theresa McCann, PhD, MPH: Development of surveys
3. Ann W. Lambert DNP, MSN,PPCNP-BC: Assistance with the quantitative arm of this study
4. Heidi Kleuss, PhD, MS: Kinesiology for the quantitative arm of this study
5. Rian Anglin, MD: Pediatric consultant
6. Mary Ann Taylor, PhD, MS: Psychological counseling
7. Leondria Hinds, DO: Survey transcribing
8. Shannon Hart, DO: Survey transcribing
9. Crystal Smith, MS: Survey transcribing
10. Jesse Hunt: Survey transcribing
11. Stephanie Kyeremeh: Data collection
12. Fernando Alvarez: Data collection
13. Tori Knapp: Data collection
14. Genevieve Thompson: Data collection
15. Madeleine Schultz: Data collection

---

**Figure 1:**
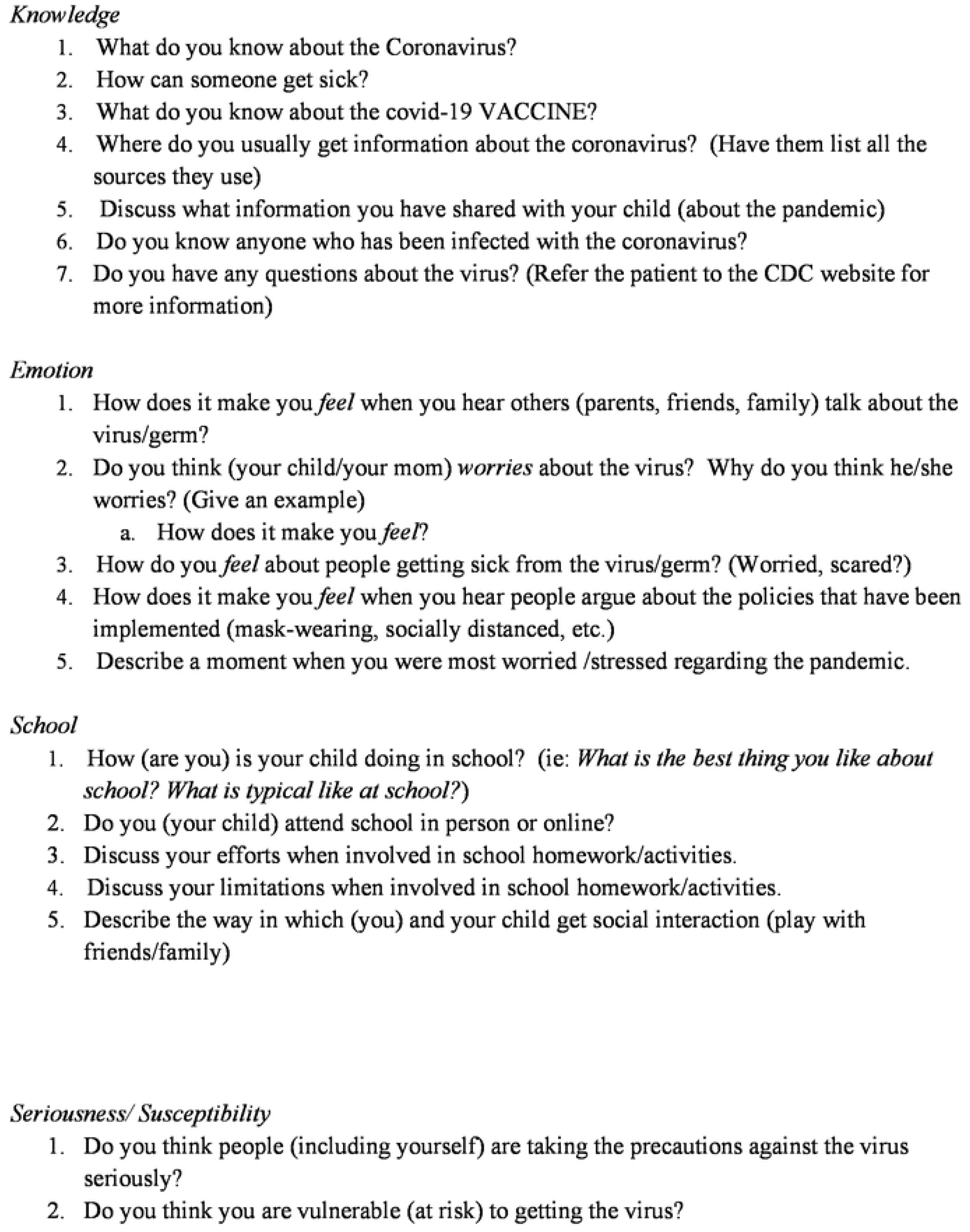
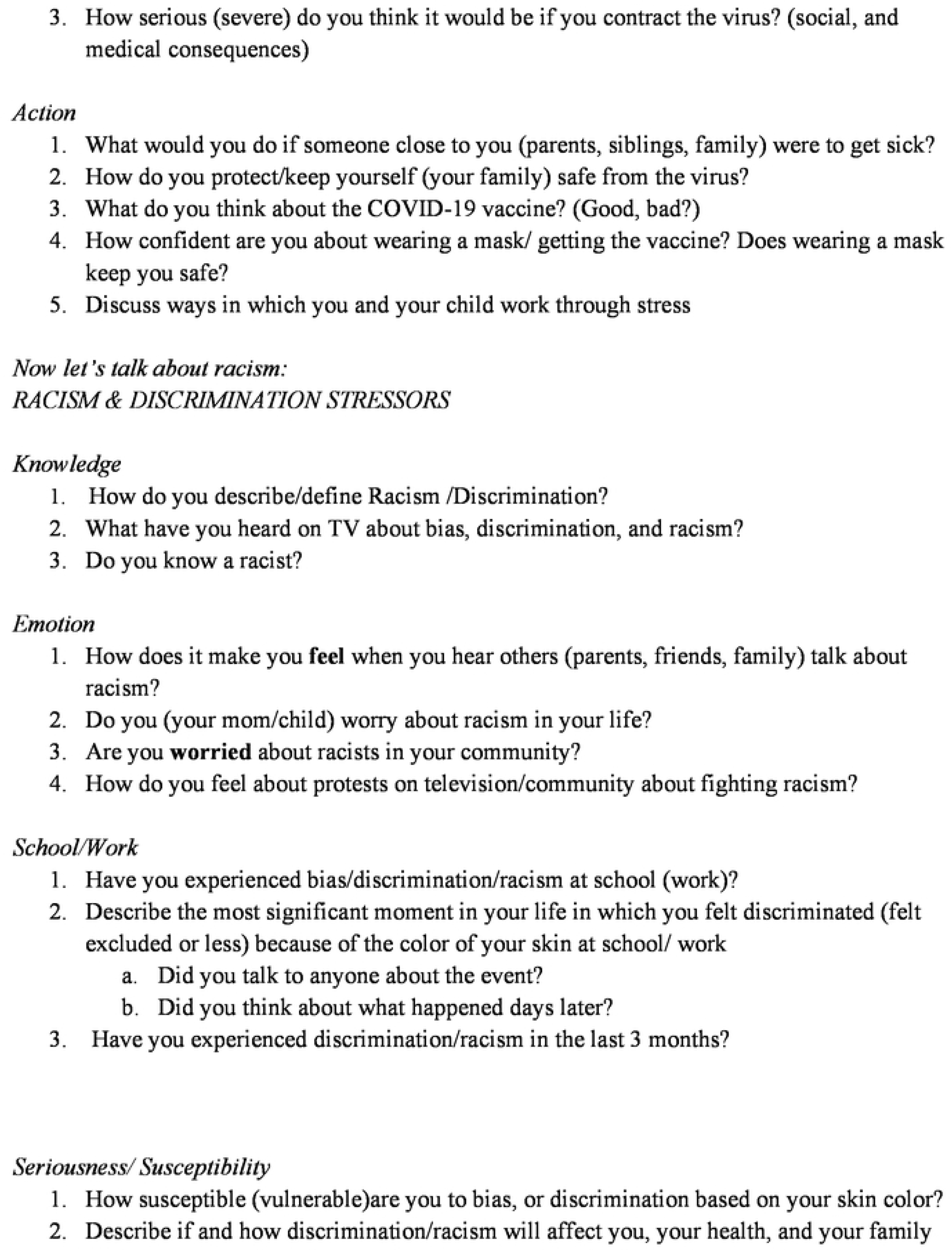
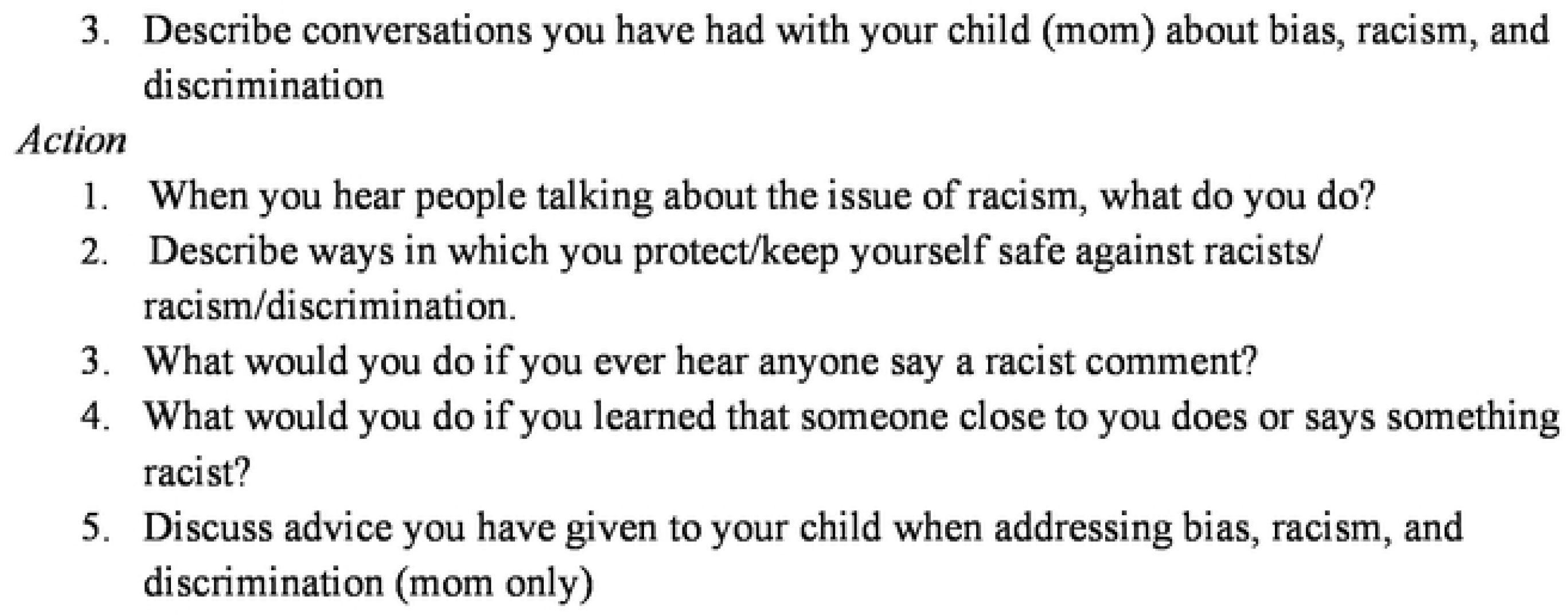
Questionnaire.

